# Contributions of Pain Interference and Affect to Patient-Reported Opioid Benefit in Chronic Pain Management

**DOI:** 10.1101/2022.12.28.22284000

**Authors:** Anne K. Baker, Su Hyoun Park, Morgan A. Rosser, Meghna Nanda, Katherine T. Martucci

**Author notes:** **Correspondence:** Katherine T. Martucci, PhD; Director, Human Affect and Pain Neuroscience Lab, Department of Anesthesiology, Duke University Medical Center, Box DUMC 3094, Durham, NC 27710 USA;, Anne K. Baker, PhD; Postdoctoral Associate, Human Affect and Pain Neuroscience Lab, Department of Anesthesiology, Duke University Medical Center, Box DUMC 3094, Durham, NC 27710 USA. **Financial Disclosures:** This project was funded by the National Institutes of Health, National Institute of Drug Abuse (NIDA), K99/R00 DA040154 and R01 DA055850 (awarded to K.T.M.). **Author Contributions:** A.K.B., and K.T.M. developed and designed the study. A.K.B., S.H.P., M.N., and K.T.M. collected and organized the data. M.A.R. provided biostatistical expertise for data analyses, and A.K.B., and M.A.R. analyzed the data. A.K.B. wrote the first draft of the manuscript. A.K.B., S.H.P, M.A.R, M.N. and K.T.M. edited and revised the manuscript.

## Abstract

**Background:** Despite known deleterious consequences associated with long-term opioid use, many individuals with chronic pain assert opioid benefits and advocate for continued opioid use. However, relative to non-opioid using chronic pain patients, opioid-using patients typically report greater pain severity and depression. Moreover, there appears to be no significant association between pain severity or interference and perceived opioid benefit among chronic pain patients. Thus, pain reduction itself might not directly relate to patients’ perceptions of opioid benefit. Given extensive prior research revealing significant overlaps between pain and affect, it is prudent to examine contributions of affective disturbances—alongside pain-related factors—to perceived opioid benefits. In the present study, we examined the hierarchical contributions of pain interference and positive affect in predicting self-reported opioid benefit. We hypothesized that positive affect combined with pain interference would best predict opioid benefit.

**Methods:** We examined multisite, cross-sectional data collected from females with fibromyalgia who were using opioids long-term (n = 40) and who were not regularly using opioids but had used them acutely (< 30 days) at least once previously (n = 25). Patients completed a set of questionnaires, including the Positive and Negative Affect Schedule, the Brief Pain Inventory, and a novel measure querying perceived opioid benefit on a 0-10 Likert scale (0 = not at all, 10 = completely). We examined relationships between pain interference, positive affect, and patient-reported opioid benefit using logistic regression.

**Results:** Among opioid-using patients, pain interference combined with positive affect was a better model for opioid benefit (AIC = 52.15) compared to pain interference alone (AIC = 57.80). However, among non-opioid using patients, pain interference alone was a better model for opioid benefit (AIC = 28.00) than pain interference combined with positive affect (AIC = 28.12).

**Conclusions:** Among patients using opioids long-term, affective factors may be primary drivers of perceived opioid benefit. Positive affect combined with pain interference modeled opioid benefit better than pain interference alone among opioid-using chronic pain patients, but not among non-opioid-using chronic pain patients. Importantly, post-hoc analyses examining the contributions of negative affect further validated the main findings; positive affect out-performed negative affect in all models. Thus, perceived opioid benefit may be a function of cumulative opioid-induced enhancements in positive affect. Based on these results, examination of factors besides pain reduction may be critical to understanding perceived opioid benefit among chronic pain patients; this understanding is essential for development of effective, opioid-sparing treatments.

**Key Points Summary:** *Question:* Does the combination of pain and affect predict patient-reported (i.e., perceived) opioid benefit better than pain alone, and do these relationships differ between groups of opioid-using and non-opioid using individuals with chronic pain?

*Findings:* Compared to pain interference alone, pain interference combined with positive affect was a better model of perceived opioid benefit among opioid-using chronic pain patients, but not among non-opioid using chronic pain patients, who had used opioids acutely at least once previously.

*Meaning:* Among patients with chronic pain using opioid medications long-term, perceived opioid benefit appears to be a consequence of opioid-related enhancements in positive affect rather than pain reduction per se.

## Introduction

Opioid misuse is a public health epidemic in the United States. While heightened clinical urgency has been aimed at addressing this crisis,^1–3^ prescription opioids for pain management remain a central part of both acute and long-term patient care, contributing to high rates of misuse, overdose, and drug-related death.^4,5^ Despite these consequences, many patients with chronic pain continue to assert opioid benefits and advocate for their use.^6^ Clearer understanding of perceived opioid benefits is thus critically needed to improve pain management, combat the opioid crisis, and promote public health.

Many studies have examined the analgesic efficacy of opioids using quantitative sensory testing,^7–9^ neuroimaging,^10–12^ and psychological questionnaires.^7^ However, these approaches to examining opioid utility have largely operated from an inadvertent assumption that perceived opioid benefit inherently stems from pain reduction. Meanwhile, very few studies have directly queried chronic pain patients’ subjective perceptions about whether and how much they benefit from opioid use. Moreover, no studies have yet empirically examined factors contributing to perceived opioid benefit.

It is logical to presume perceived opioid benefit is primarily driven by pain reduction. However, as revealed by limited prior research that includes patient perspectives, opioid benefit is not significantly associated with pain severity or pain interference.^6^ Furthermore, opioid-using chronic pain patients paradoxically report greater pain severity than patients who do not use opioids,^6^ suggesting pain reduction is not the sole required factor for perceived opioid benefit.

Given extensive clinical and neurobiological overlap between pain and affect,^13–15^ affective factors may be integral to perceived opioid benefit. Patients with chronic pain exhibit high rates of affective and mood disturbances which, in turn, are associated with opioid craving and risk for opioid misuse.^16–18^ Positive affect, in particular, appears to be a critical mechanism linking chronic pain and opioid use.^19^ As proposed previously, positive affect may be a key pain inhibitory factor that, when blunted in chronic pain patients, increases opioid use.^20^ While perceived opioid benefit and opioid use are not the same thing, it is reasonable to expect that continued opioid use follows from at least some degree of perceived benefit. Thus, affective factors relating to opioid use may also relate to perceived opioid benefit.

Accordingly, we examined contributions of both pain interference and positive affect to self-reported perceptions of opioid benefit. We hypothesized the combination of positive affect and pain interference would predict opioid benefit better than pain interference alone.

## Methods

### Participants

Data were obtained from females with fibromyalgia (N = 70) at Stanford University (non-opioid using patients with fibromyalgia, FMN, n =10; opioid-using patients with fibromyalgia, FMO, n=17) and at Duke University (FMN n= 18; FMO n = 25). Five participants were excluded from analyses due to missing data (**Figure 1**), resulting in 65 participants with complete data for analysis (FMN n = 25, FMO n = 40). All participants were required to meet the modified American College of Rheumatology 2011 criteria for fibromyalgia, which included (1) a widespread pain index (WPI) plus a symptom severity (SS) score ≥ 5, or WPI score 3-6 plus SS score ≥ 9, (2) comparable symptoms present for at least 3 months, and (3) no diagnosis that would otherwise explain the pain. Additionally, participants were required to have pain in all 4 body quadrants, an average 0-10 verbal pain scale rating ≥ 2, and no uncontrolled psychiatric disorders. All FMN participants had not taken any opioid medications in the 90 days prior to the study visit and their total lifetime opioid medication usage was < 30 days. All FMO participants were required to have taken opioid medications regularly for at least the past 90 days prior to the study visit. FMO participants’ opioid doses were transformed into daily morphine milligram equivalents (MME; see **Table 1**). Non-opioid medication use (e.g., anti-depressants, non-steroidal anti-inflammatory medications) is reported in **Table 2**.

**Table 1.** Group Differences in Participant Characteristics

|  | FMO |  | FMN |  | t | p |
| --- | --- | --- | --- | --- | --- | --- |
|  | Mean | SD | Mean | SD |  |  |
| Morphine Milligram Equivalents | 30.73 | 54.70 |  |  |  |  |
| Opioid Use Duration (Years) | 7.24 | 9.15 |  |  |  |  |
| Age | 50.95 | 8.70 | 41.08 | 13.24 | -4.33 | <0.001*** |
| Opioid Benefit | 7.93 | 2.03 | 5.79 | 3.26 | -3.10 | 0.003** |
| Opioid-induced Pain Reduction | 2.26 | 3.17 | 2.10 | 4.05 | -0.18 | 0.86 |
| Non-opioid analgesic-induced Pain Reduction | 0.82 | 2.07 | 1.55 | 2.3 | 1.51 | 0.14 |
| BPI Pain Severity | 5.86 | 1.35 | 5.53 | 1.53 | -1.10 | 0.28 |
| BPI Pain Interference | 6.38 | 2.21 | 6.59 | 2.34 | 0.46 | 0.65 |
| PANAS Positive Affect | 25.23 | 7.33 | 24.75 | 7.58 | -0.31 | 0.75 |
| PANAS Negative Affect | 20.55 | 7.38 | 21.20 | 7.07 | 0.44 | 0.66 |
| STAI State Anxiety | 41.31 | 12.58 | 40.65 | 8.79 | -0.30 | 0.76 |
| STAI Trait Anxiety | 46.49 | 11.62 | 46.98 | 9.54 | 0.23 | 0.82 |
| POMS Total Mood Disturbance | 24.34 | 18.66 | 22.90 | 16.56 | -0.40 | 0.69 |

Group differences in participant characteristics were conducted via independent-samples t-tests. \*\*denotes $p < 0.01$ , \*\*\* $p < 0.001$ . BPI = Brief Pain Inventory, PANAS = Positive and Negative Affect Schedule, STAI = State-Trait Anxiety Inventory, POMS = Profile of Mood States.

**Table 2.** Non-opioid Pain and Mood-Altering Medication Use Frequencies

| Medication | FMO (n) | FMN (n) |
| --- | --- | --- |
| Tapentadol | 0 | 2 |
| Tramadol | 0 | 16 |
| NSAIDs | 10 | 21 |
| Acetaminophen | 4 | 22 |
| Other Pain Meds (e.g., lidocaine) | 2 | 1 |
| SNRIs | 8 | 15 |
| SSRIs | 5 | 8 |
| Tricyclic Antidepressants | 3 | 2 |
| Other Anxiolytics | 3 | 5 |
| Triptans | 3 | 8 |
| Other SRIs | 1 | 7 |
| NDRIs | 1 | 5 |
| SR Antagonists | 1 | 6 |
| Antiepileptics | 4 | 10 |
| Benzodiazepines & Benzodiazepine-like | 7 | 13 |
| Muscle Relaxants | 6 | 16 |
| GABA Analogs | 6 | 16 |
| Birth Control | 7 | 7 |

Participants self-reported regular use of any pain and/or mood-altering medications. Qualitative responses were transformed into binary yes (1) – no (0) responses and frequencies were computed for all yes answers in each substance category. NSAID = non-steroidal anti-inflammatory, SNRI = selective norepinephrine reuptake inhibitor, SSRI = selective serotonin reuptake inhibitor, SRI = serotonin reuptake inhibitor, NDRI = norepinephrine-dopamine reuptake inhibitor, SR = serotonin receptor, GABA = $\gamma$ -Aminobutyric acid.

**Figure 1.**
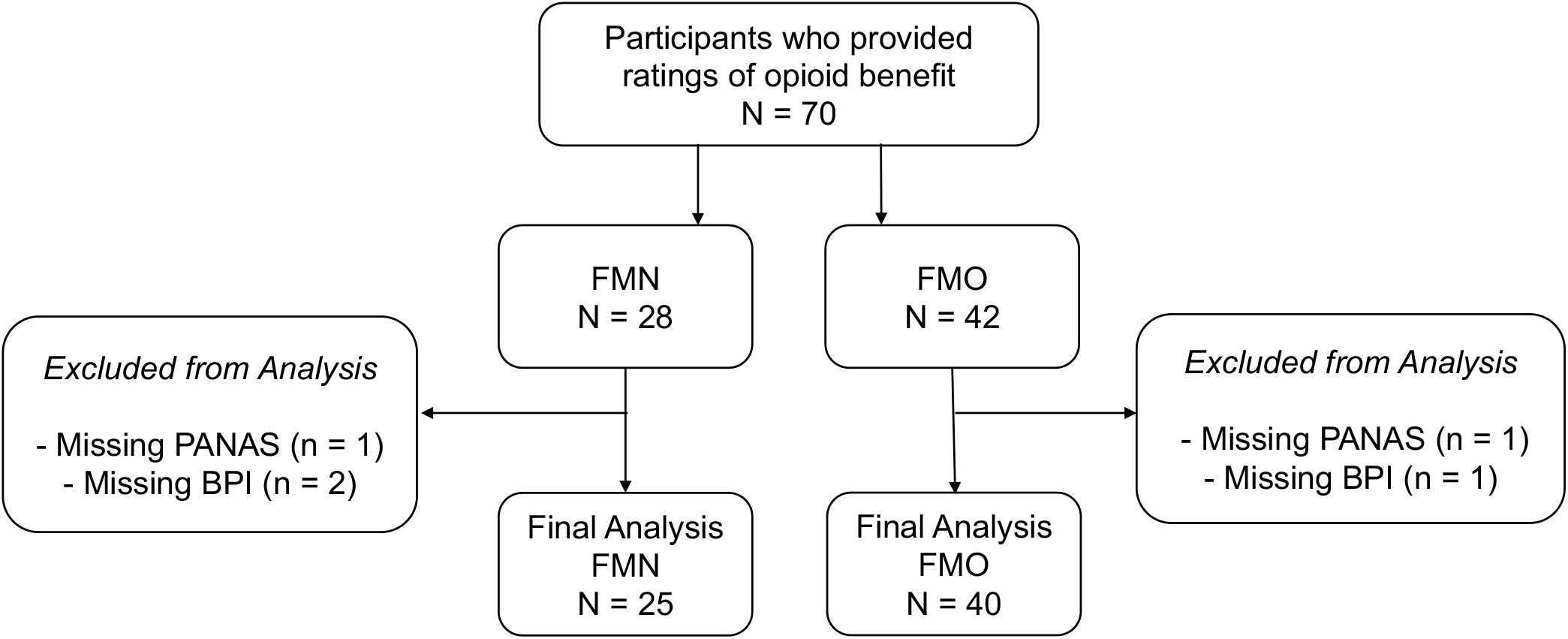
Flow chart of participant inclusions and exclusions for the analysis. FMN, non-opioid using patients with fibromyalgia (i.e., retrospective ratings of opioid benefit); FMO, opioid-using patients with fibromyalgia; PANAS = Positive and Negative Affect Schedule, BPI = Brief Pain Inventory.

### Procedures

Study participants completed a battery of standard clinical questionnaires, which included the Brief Pain Inventory Short Form (BPI),^21^ the Positive and Negative Affect Schedule (PANAS),^22^ the Profile of Mood States (POMS),^24^ and the State-Trait Anxiety Inventory (STAI).^25^ Participants also completed a novel self-report measure (see Appendix A) querying (1) perceived opioid and non-opioid analgesic (e.g., ibuprofen) medication benefits, (2) average pain while taking opioid and non-opioid analgesic medications, and (3) average pain when *not* taking opioid and non-opioid analgesic medications. Medication-induced pain reduction was computed by subtracting average pain when taking opioid/non-opioid medications from average pain when not taking opioid/non-opioid medications. Participants who were not taking opioid medications at the time of the study (FMN participants) were instructed to retrospectively rate opioid benefit and opioid-induced pain reduction by recalling prior lifetime experience(s) of taking opioids. All participants completed written informed consent prior to data collection and all study procedures were approved by the Stanford University and Duke University Institutional Review Boards. An IRB-approved Data Use Agreement was created to allow for the data collected at Stanford University to be analyzed by the research team at Duke University.

### Statistical Analysis

Statistical analyses were conducted using IBM SPSS 26 (IBM Corporation, Armonk, NY, USA) and RStudio 4.1.3.^26^ Spearman’s correlations were conducted to examine relationships between opioid benefit and all 4 pain- and affect-related variables (BPI, PANAS, POMS, STAI), as well as morphine dosage, pain duration, and age. Group differences between FMO and FMN participants were evaluated with an independent samples t-test. To assess the contributions of pain interference and positive affect to perceived opioid benefit, planned analyses were nested linear regressions stratified by group. However, Shapiro-Wilk tests revealed that both FMOs (p = 0.024) and FMNs (p = 0.029) failed to meet the required assumption of normally distributed residuals. Transformations of the data allowed for FMNs to meet this assumption, but FMOs still failed to meet this assumption after data transformations. Thus, for consistency, we proceeded with logistic regression for both groups. To allow for logistic regression, the perceived opioid benefit outcome variable was converted to a binary variable using the median score of 7 as the cut-point. For each group, we used Akaike’s information criteria (AIC)^28^ for comparisons between models. The models considered were: (1) the main effect of pain interference on perceived opioid benefit, (2) the main effects of pain interference and positive affect on perceived opioid benefit, and (3) the main effects of pain interference, positive affect, and the interaction between the two on perceived opioid benefit. All models met the goodness of fit criteria according to the Hosmer and Lemeshow test or an equivalent goodness of fit test using the R package “rms”. We also conducted exploratory post-hoc analyses comparing positive affect to a range of negative affective variables (see additional methods described in the Supplemental Materials).

## Results

Group differences in demographic and clinical variables were assessed between patients currently taking opioids (FMO) and patients who were not currently taking opioids but who had taken them acutely (i.e., for a period of less than 30 days) previously (FMN). Significant group differences in age were observed (t = −4.33, *p* < 0.001), revealing FMO patients were significantly younger than FMN patients. Significant differences were also observed in perceived opioid benefit (t = −3.10, *p* = 0.002), with FMO patients endorsing significantly greater opioid benefit than FMN patients. No significant differences in any affective variable, average pain severity or intensity, or pain duration were observed between the two groups (**Table 1**).

Spearman’s correlations revealed opioid benefit was not significantly correlated with average opioid-induced pain reduction (*ρ* = 0.06, *p* = 0.70) among FMO participants (**Figure 2a**). In contrast, opioid benefit was significantly positively correlated with average retrospectively rated opioid-induced pain reduction during acute prior opioid use among FMN participants (*ρ* = 0.41, *p* = 0.04). For self-reported non-opioid analgesic medication benefit (i.e., acetaminophen, ibuprofen), we observed no significant associations with average medication-induced pain reduction among FMO participants (*ρ* = 0.11, *p* = 0.53), or among FMN participants (*ρ* = 0.17, *p* = 0.25) (**Figure 2b**).

**Figure 2.**
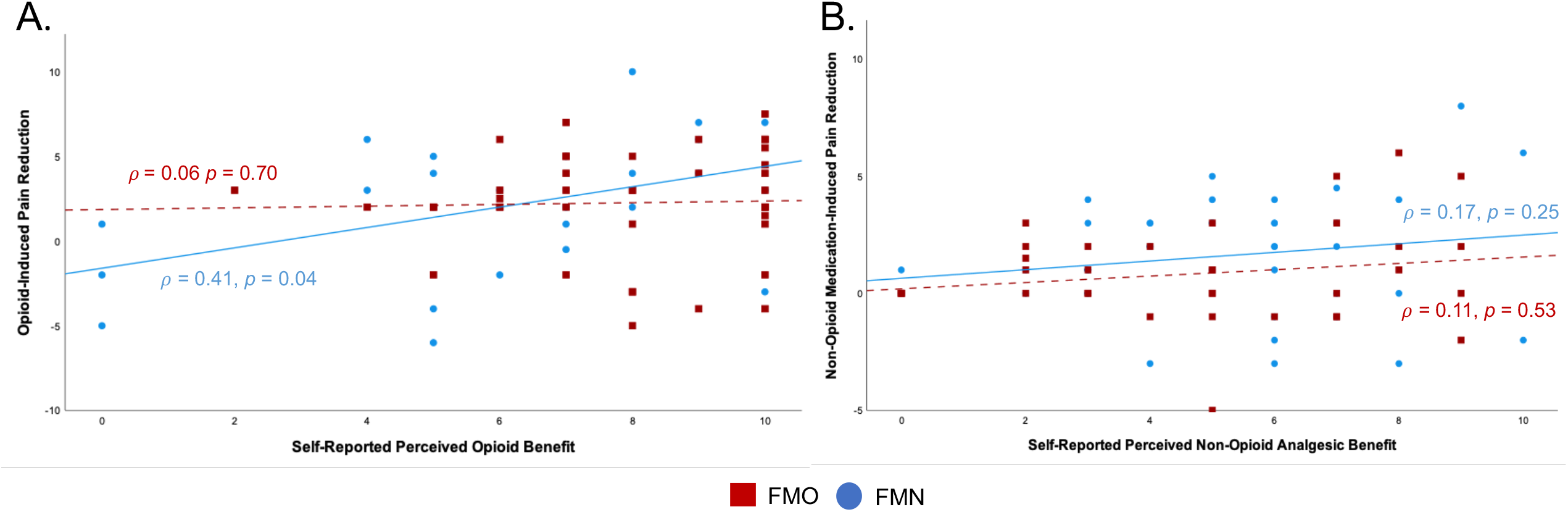
A) Spearman correlations between perceived opioid benefit and opioid-induced pain reduction are represented separately for opioid-using patients (FMO, red) and non-opioid using patients (FMN, blue). B) Spearman correlations between perceived non-opioid analgesic medication benefit and non-opioid medication-induced pain reduction are represented separately for opioid-using patients (FMO, red) and non-opioid using patients (FMN, blue). Spearman correlations were exploratory and thus not corrected for multiple comparisons.

Among FMO participants, opioid benefit was positively correlated with pain interference (*ρ* = 0.32, *p* = 0.05), and was not significantly associated with any affective measure, opioid dose (in morphine milligram equivalents (MME)), opioid use duration, pain duration, or pain severity. Pain interference was positively associated with negative affect (*ρ* = 0.42, *p* = 0.005), mood disturbance (*ρ* = 0.59, *p* < 0.001), state anxiety (*ρ* = 0.50, *p* < 0.001), trait anxiety (*ρ* = 0.40, *p* = 0.008), and pain severity (*ρ* = 0.48, *p* < 0.001). Pain interference was also inversely associated with positive affect (*ρ* = −0.36, *p* = 0.02). Although significant correlations were identified between the predictors of our models, evaluation of the variance inflation factor (VIF) ruled out collinearity as a potential issue.

Among FMN participants, opioid benefit was significantly inversely associated with pain interference (*ρ* = −0.53, *p* = 0.006); no significant correlations were observed between opioid benefit and any affective measure, pain severity, or pain duration. Pain interference was positively associated with mood disturbance (*ρ* = 0.50, *p* < 0.001), state anxiety (*ρ* = 0.40, *p* = 0.005), and pain severity (*ρ* = 0.38, *p* = 0.008). The evaluation of VIF also ruled out collinearity as a potential issue in the FMN cohort. All Spearman correlations are reported in **Table 2 in the Supplemental Materials**.

Among FMO participants, the logistic regression model with the main effects of both positive affect and pain interference was the best model for predicting opioid benefit (AIC = 52.15, *p* = 0.006); the comparison models were pain interference alone (AIC = 57.80), and both main effects plus the interaction between the two (AIC = 52.88, *p* = 0.262). Among FMNs, the model of best fit predicting opioid benefit was pain interference alone (AIC = 28.00); the comparison models were the main effects of pain interference and positive affect (AIC = 28.12, *p* = 0.167), and both main effects plus the interaction between the two (AIC = 28.88, *p* = 0.210). Complete logistic regression results for both groups are presented in **Table 3**.

**Table 3.** Logistic Regression Models of Perceived Opioid Benefit

| Model | FMO |  |  | FMN |  |  |
| --- | --- | --- | --- | --- | --- | --- |
|  | *Odds Ratio (95% CI)* | *p-value* | *AIC* | *Odds Ratio (95% CI)* | *p-value* | *AIC* |
| <u>Base</u> |  |  | 57.80 |  |  | **28.00** |
| Pain Interference | 1.16 (0.82, 1.62) | 0.40 |  | 0.49 (0.25, 0.94) | 0.03 |  |
| <u>Main Effects</u> |  |  | **52.15** |  |  | 28.12 |
| Pain Interference | 1.48 (0.96, 2.28) | 0.08 |  | 0.54 (0.29, 1.02) | 0.06 |  |
| Positive Affect | 1.16 (1.03, 1.31) | 0.01 |  | 1.11 (0.95, 1.29) | 0.20 |  |
| <u>Main Effects + Interaction</u> |  |  | 52.88 |  |  | 28.88 |
| Pain Interference | 2.75 (0.80, 9.46) | 0.11 |  | 0.06 (0.00, 1.86) | 0.21 |  |
| Positive Affect | 1.40 (0.97, 2.02) | 0.07 |  | 0.64 (0.22, 1.82) | 0.40 |  |
| Pain Interference *Positive Affect | 0.97 (0.92, 1.02) | 0.28 |  | 1.09 (0.93, 1.28) | 0.31 |  |

Contributions of pain interference and positive affect to perceived opioid benefit were examined with logistic regression. Akaike's information criteria (AIC) was used to compare models. FMO = opioid-using fibromyalgia patients; FMN = non-opioid using fibromyalgia patients

In post-hoc exploratory analyses stratified by cohort, we evaluated main effects models of pain interference combined with negative affect, state anxiety, trait anxiety, and mood disturbance. Compared to these exploratory models, the original models remained the best fit for each group (i.e., main effect of pain interference alone for FMN, main effects of pain interference and positive affect for FMO). All results from the exploratory model analyses are reported in the **Supplemental Materials**.

## Discussion

For the first time, this study evaluated how pain and affective factors, both separately and in combination with one another, contribute to patient-reported (i.e., perceived) opioid benefit. In a sample of females with fibromyalgia who were using opioids long-term (i.e., FMOs), the combined main effects of pain interference and positive affect modeled opioid benefit (1) better than pain interference alone, and (2) better than the interaction between pain interference and positive affect. These results are consistent with limited prior research, which demonstrates discrepancies between clinical outcomes and patient perspectives of opioid benefits.^6^ Together, these studies highlight the importance of evaluating factors beyond pain reduction that may influence chronic pain patients’ perceptions of opioid benefit.

We further evaluated contributors to opioid benefit within a comparison sample: non-opioid using females with fibromyalgia (FMNs) who had acutely (< 30 days) used opioids at least once previously. In contrast to FMOs, opioid benefit among FMNs was better predicted by pain interference alone than by the combination of pain interference and positive affect. Given that inclusion in our cohorts was defined by opioid use, the observed differential contributions of pain and affect to perceived opioid benefit may reflect the effects of long-term opioid use among chronic pain patients. Additionally, as our participants were self-selected for opioid vs. non-opioid use prior to their participation, other variables due to different medication preferences, physiological responses to medication use, and/or severity of clinical condition could have contributed to our observed group differences.

We hypothesized that perceived opioid benefit would be observed as a function of positive affect combined with pain interference. In line with our hypothesis, we observed that high positive affect combined with high pain interference best predicted greater opioid benefit among long-term opioid users. As higher positive affect was associated with greater opioid benefit among long-term opioid users, our results align with the theoretical perspective that opioids may offset blunted positive affect. Blunted positive affect is commonly observed in patients with chronic pain,^27–29^ and exogenous opioids may serve to temporarily ameliorate this deficiency.^20^ However, long-term opioid use may also contribute to blunted positive affect through dysregulation of mesolimbic reward circuitry.^30^ Indeed, mesolimbic reward circuits are altered in individuals with chronic pain^31,32^, including fibromyalgia.^33–35^ Additional changes to reward circuits have been observed in patients with chronic pain who take opioid medications long-term.^36,37^

As the results from this study were derived from cross-sectional data, it is possible that we could have captured one point of an allostatic process of opioid use that, if continued, may paradoxically yield decreased positive affect and, ultimately, less perceived benefit over time. Nevertheless, as suggested by our results in the context of long-term opioid use, patient-reported (i.e., perceived) opioid benefit may reflect cumulative opioid-related enhancements in positive affect for at least some period of time. Further, positive affect may be a uniquely important contributor to perceived opioid benefit given that positive affect out-performed negative affect in all models of opioid benefit.

The parallel analyses among FMNs and FMOs provided contrasting yet compelling results. While FMO-reported opioid benefit was best modeled by pain interference combined with positive affect, FMN-reported (i.e., retrospective) perceived opioid benefit was best modeled by pain interference alone. These contrasting results between our cohorts provide evidence that positive affect is an essential feature of perceived opioid benefit, exclusively in the context of long-term opioid therapy in patients with chronic pain.

We additionally analyzed relationships between perceived opioid benefit and average pain ratings among our two cohorts. Among FMOs, opioid benefit was not significantly related to average pain while taking opioids. By contrast, among FMNs, opioid benefit was significantly inversely related to average pain while taking opioids. These group differences complement the model results discussed above. Together, these data support the conclusion that pain reduction is not a sole contributor to opioid benefit among long-term opioid users. Moreover, because pain reduction did predict retrospectively rated opioid benefit (i.e., during past acute opioid use), it is possible that the condition of long-term opioid use changes the relationship between pain and opioid benefit.

Several limitations should be considered. First, the cross-sectional nature of the data imparts directional limitations. Future research will require longitudinal, prospective studies initiated at commencement of opioid therapy; such studies could examine the extent to which pre-opioid levels of positive affect and pain interference contribute to perceived opioid benefit over time. We also note potential retrospective bias among FMNs who rated opioid benefit by recalling previous experiences of acute opioid use. Further, we were unable to control for variability due to potential neurophysiologic differences between acute vs. long-term opioid use. Indeed, long-term opioid use may significantly influence the relationship between positive affect and perceived opioid benefit. Additionally, while superiority of one model was determined over the others, model estimates yielded high variability. As such, future studies should consider increasing the sample population to decrease this variability and to thereby test these models more effectively. Further, despite adequate power for the present analyses as determined per our power analysis, our modest sample size prevented us from additionally controlling for menstrual cycle phase and hormonal factors (e.g., birth control). Given well established differences in pain and affect across the menstrual cycle,^38–41^ menstrual cycle-related variability is a vital consideration for future research aimed to characterize contributors to perceived opioid benefit. Finally, the perceived opioid benefit questionnaire is not yet a validated measure; psychometric studies verifying the internal consistency and test-retest reliability of this novel measure will provide an asset to future research examining opioid benefits.

Overall, the present study contributes novel and important findings to the existing knowledge on perceived opioid benefit among individuals with chronic pain. In summary, our results in long-term opioid-using chronic pain patients indicated that positive affect combined with pain interference predicts perceived opioid benefit better than pain interference alone. We posit that positive affect may serve as part of a feedback loop that links chronic pain to perceived opioid benefit and, theoretically, also to continued opioid use (**Figure 3**). Future longitudinal studies will be essential to further evaluate these relationships.

**Figure 3.**
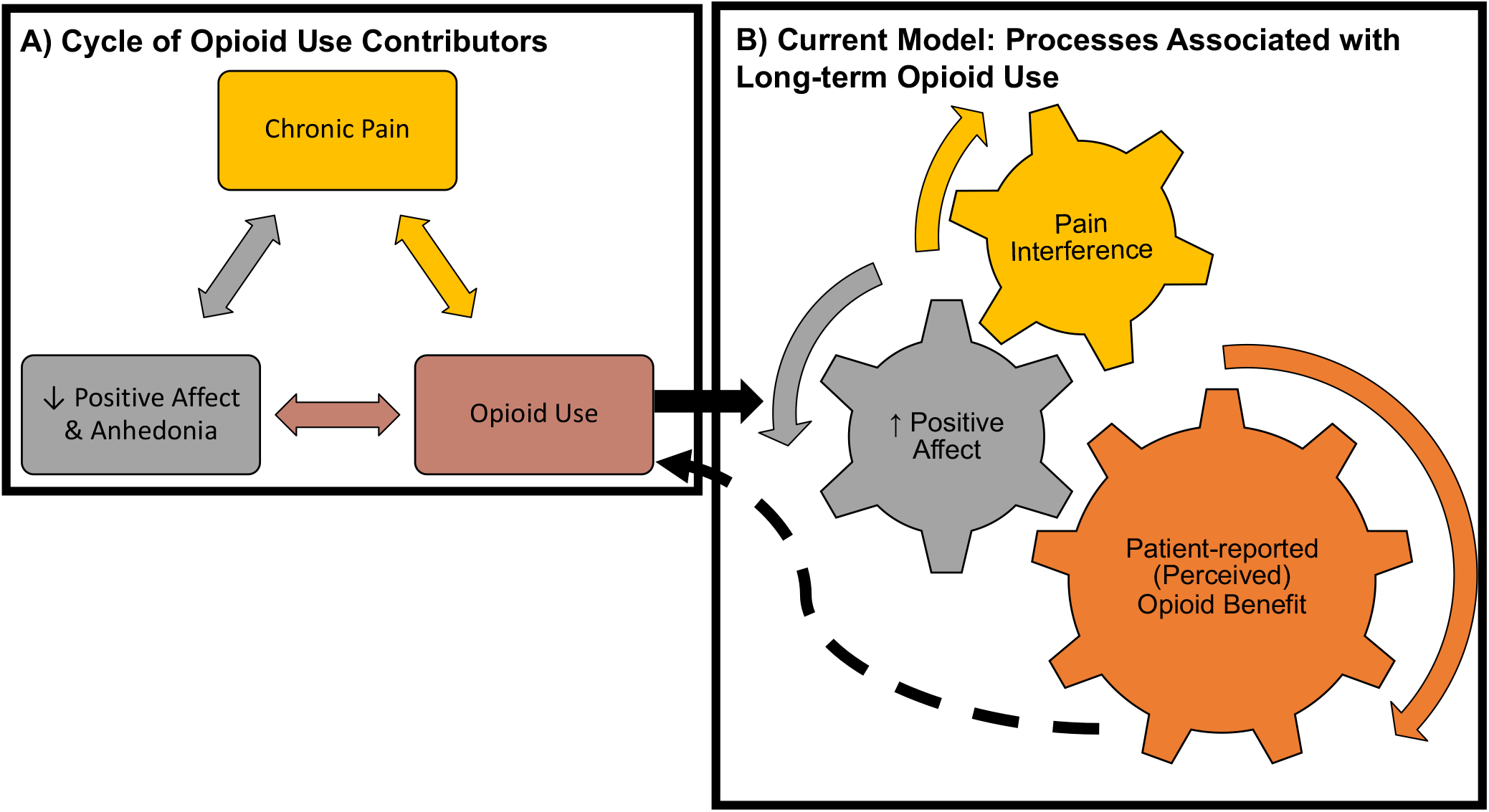
**Panel A** represents an established set of bidirectional relationships between chronic pain, low positive affect (including its most extreme form, anhedonia), and opioid use. **Panel B** represents the cross-sectional results from the present study. By itself, pain interference does not significantly impact self-reported (i.,e., perceived) opioid benefit among chronic pain patients using opioids long-term. As such, the gears for pain interference and opioid benefit do not directly interact. Instead, the addition of positive affect into the model is what drives perceived opioid benefit. The black arrows represent a hypothesized longitudinal model in which our present results join with current theory in a positive feedback loop. The solid arrow moving from panel A to B represents the euphoric effects of opioids that may increase positive affect. The dashed arrow represents a hypothesis that greater perceived opioid benefit contributes to the continuation of long-term opioid therapy in patients.

## Data Availability

All data produced in the present study are available upon reasonable request to the authors.

## Acknowledgements

We thank Lindsie Boerger, Dr. Cristina (Cojocaru) Mackey, and Erin Perrine for their assistance with participant recruitment and data collection. Special thanks also to Dr. Sean C. Mackey for additional support on the portion of the study data collected at Stanford University. We especially thank all the participants for their generosity of time and efforts spent in this study.

