## Supplemental Materials for "Contributions of Pain Interference and Affect to Patient-Reported Opioid Benefit in Chronic Pain Management"

**Methods**

**Statistical Analysis**

Post-hoc exploratory statistical analyses were conducted using R version 4.1.3. To further assess the unique contributions of positive affect to perceived opioid benefit, we also evaluated the contributions of a range of negative affective variables to opioid benefit. We compared 6 models: 1) the main effect of pain interference, 2) the main effects of pain interference and positive affect, 3) the main effects of pain interference and negative affect, 4) the main effects of pain interference and state anxiety, 5) the main effects of pain interference and trait anxiety, and 6) the main effects of pain interference and mood disturbance.

As with our primary analysis, we used logistic regression and converted our outcome using the median score of 7 for a cutoff. The new opioid benefit outcome was then defined as a binary term, with 1 representing an opioid benefit score greater than 7, and 0 representing an opioid benefit score less than or equal to 7. Analyses were stratified by group (opioid-using fibromyalgia patients (FMO) and non-opioid using fibromyalgia patients (FMN)), and for each group we used Akaike’s information criteria (AIC)^28^ to compare model fit. Because we did not find any interaction effects in the primary analyses, we only examined main effects.

**Results**

In the FMO cohort, the logistic regression model with the main effects of both positive affect and pain interference performed as the best model for predicting opioid benefit (AIC = 52.15), relative to the models with 1) the main effect of pain interference (AIC = 57.80), 2) the main effects of pain interference and negative affect (AIC = 57.03), 3) the main effects of pain interference and state anxiety (AIC = 59.17), and 4) the main effects of pain interference and mood disturbance (AIC = 57.67) (Supplemental Table 1). In the FMO cohort, the model with the main effects of pain interference and trait anxiety did not pass the Hosmer and Lemeshow goodness of fit test used to assess goodness of fit in logistic models; accordingly, we have not reported values for this model.

Among FMNs, the logistic regression model with the main effect of pain interference alone performed as the best model for predicting opioid benefit (AIC = 28.00), relative to the models with 1) the main effects of pain interference and positive affect (AIC = 28.12), 2) the main effects of pain interference and negative affect (AIC = 29.11), 3) the main effects of pain interference and state anxiety (AIC = 29.87), 4) the main effects of pain interference and trait anxiety (AIC = 29.05), and 5) the main effects of pain interference and mood disturbance (AIC = 57.67) (Supplemental Table 1).

| Supplemental Table 1. Comparison of Logistic Regression Models Predicting Perceived Opioid Benefit | | | | | |
| --- | --- | --- | --- | --- | --- |
| **Model** | **Pain Interference** | | **Covariate** | | ***AIC*** |
|  | ***Odds Ratio (95% CI)*** | ***p-value*** | ***Odds Ratio (95% CI)*** | ***p-value*** |  |
| FMO |  |  |  |  |  |
| Pain Interference | 1.16 (0.82, 1.62) | 0.400 | -- | -- | 57.80 |
| Pain Interference + Positive Affect | 1.48 (0.96, 2.28) | 0.075 | 1.16 (1.03, 1.31) | 0.014 | **52.15** |
| Pain Interference + Negative Affect | 1.37 (0.89, 2.10) | 0.147 | 0.92 (0.82, 1.02) | 0.112 | 57.03 |
| Pain Interference + State Anxiety | 1.25 (0.84, 1.85) | 0.268 | 0.97 (0.91, 1.04) | 0.433 | 59.17 |
| Pain Interference + Trait Anxiety | ***--*** | -- | -- | -- | -- |
| Pain Interference + Mood Disturbance | 1.41 (0.90, 2.20) | 0.130 | 0.97 (0.92, 1.01) | 0.157 | 57.67 |
| FMN |  |  |  |  |  |
| Pain Interference | 0.49 (0.25, 0.94) | 0.032 | **--** | **--** | **28.00** |
| Pain Interference + Positive Affect | 0.54 (0.29, 1.02) | 0.056 | 1.11 (0.95, 1.29) | 0.200 | 28.12 |
| Pain Interference + Negative Affect | 0.48 (0.25, 0.92) | 0.027 | 1.07 (0.92, 1.25) | 0.361 | 29.11 |
| Pain Interference + State Anxiety | 0.46 (0.22, 0.96) | 0.038 | 1.02 (0.90, 1.16) | 0.714 | 29.87 |
| Pain Interference + Trait Anxiety | 0.44 (0.21, 0.94) | 0.034 | 1.05 (0.95, 1.17) | 0.339 | 29.05 |
| Pain Interference + Mood Disturbance | 0.45 (0.21, 0.94) | 0.033 | 1.02 (0.95, 1.10) | 0.581 | 29.69 |
| Comparison of logistic regression models predicting a score higher than 7 for opioid benefit in each cohort with pain interference as a predictor and pain interference adjusting for positive affect, negative affect, state anxiety, trait anxiety, or mood disturbance. Models were Bonferroni corrected for multiple comparisons. | | | | | |

| Supplemental Table 2. Relationships Between Perceived Opioid Benefit and Clinical Measures | | | | |
| --- | --- | --- | --- | --- |
|  | **FMO** | | **FMN** | |
| **Measures** | $\boldsymbol{\rho}$ | ***p*** | $\boldsymbol{\rho}$ | ***p*** |
| Morphine Milligram Equivalents | 0.26 | 0.10 |  |  |
| Opioid Use Duration (Years) | -0.12 | 0.66 |  |  |
| Pain Duration | 0.04 | 0.82 | 0.52 | 0.12 |
| Opioid-induced Pain Reduction | 0.06 | 0.70 | 0.41 | 0.04* |
| BPI Pain Severity | 0.13 | 0.42 | -0.22 | 0.28 |
| BPI Pain Interference | 0.32 | 0.05* | -0.53 | 0.006** |
| PANAS Positive Affect | 0.28 | 0.08 | 0.32 | 0.11 |
| PANAS Negative Affect | -0.09 | 0.58 | -0.16 | 0.43 |
| STAI State Anxiety | -0.08 | 0.61 | -0.20 | 0.32 |
| STAI Trait Anxiety | -0.10 | 0.54 | -0.02 | 0.91 |
| POMS Total Mood Disturbance | -0.04 | 0.81 | -0.28 | 0.15 |
| Spearman’s Correlations were conducted to test relationships between perceived opioid benefit ratings and several clinical measures relating to pain and affect. Correlations were tested for the opioid-using (FMO) and non-opioid using patient (FMN) groups separately. *denotes p < 0.05, uncorrected **p < 0.01, uncorrected. BPI = Brief Pain Inventory, PANAS = Positive and Negative Affect Schedule, STAI = State-Trait Anxiety Inventory, POMS = Profile of Mood States. | | | | |
